# Idiopathic Intracranial Hypertension and Cardiovascular Diseases Risk in the United Kingdom Women: An Obesity-Adjusted Risk Analysis Using Indirect Standardization

**DOI:** 10.1101/2024.10.20.24315837

**Authors:** Ahmed Y. Azzam, Mahmoud M. Morsy, Mohamed Hatem Ellabban, Ahmed M. Morsy, Adham Adel Zahran, Mahmoud Nassar, Omar S. Elsayed, Adam Elswedy, Osman Elamin, Ahmed Saad Al Zomia, Hana J. Abukhadijah, Hammam A. Alotaibi, Oday Atallah, Mohammed A. Azab, Muhammed Amir Essibayi, Adam A. Dmytriw, Mohamed D. Morsy, David J. Altschul

**Affiliations:** Faculty of Medicine, October 6 University, 6^th^ of October City, Giza, Egypt; Montefiore-Einstein Cerebrovascular Research Lab, Albert Einstein College of Medicine, Bronx, NY, USA; Director of Clinical Research and Clinical Artificial Intelligence, American Society for Inclusion, Diversity, and Health Equity (ASIDE), Delaware, USA; Clinical Research Fellow, American Society for Inclusion, Diversity, and Health Equity (ASIDE), Delaware, USA; Faculty of Medicine, Al-Azhar University, Cairo, Egypt; Kasr Alainy Faculty of Medicine, Cairo University Hospitals, Cairo University, Cairo, Egypt; Department of Medicine, Division of Endocrinology, Diabetes and Metabolism, Jacobs School of Medicine and Biomedical Sciences, University at Buffalo, New York, USA; Founder, American Society for Inclusion, Diversity, and Health Equity (ASIDE), Delaware, USA; Department of Neurosurgery, Jordan Hospital, Amman, Jordan; College of Medicine, King Khalid University, Abha, Saudi Arabia; Medical Research Center, Hamad Medical Corporation, Doha, Qatar; Ophthalmology Department, Prince Sultan Military Medical City, Riyadh, Saudi Arabia; Department of Neurosurgery, Hannover Medical School, Hannover, Germany; Department of Neurosurgery, Cleveland Clinic Foundation, Cleveland, OH, USA; Department of Neurological Surgery, Montefiore Medical Center, Albert Einstein College of Medicine, Bronx, NY, USA; Neuroendovascular Program, Massachusetts General Hospital & Brigham and Women’s Hospital, Harvard University, Boston, MA, USA; Neurovascular Centre, Divisions of Therapeutic Neuroradiology & Neurosurgery, St.; Michael’s Hospital, University of Toronto, Toronto, ON, Canada. 18- College of Medicine, King Khalid University, Abha, Saudi Arabia

**Keywords:** Idiopathic Intracranial Hypertension, Pseudotumor Cerebri, Stroke, Ischemic Stroke, Cardiovascular Disease

## Abstract

**Introduction:** Idiopathic intracranial hypertension (IIH) is associated with increased cardiovascular disease (CVD) risk, but the relative contributions of obesity versus IIH-specific factors remain unclear. This study aims to disentangle the effects of obesity and IIH on stroke and CVD risk, building upon previous research suggesting a two-fold increased risk of cardiovascular events in women with IIH compared to BMI-matched controls.

**Methods:** We conducted an obesity-adjusted risk analysis using Indirect Standardization analysis based on Adderley et al. study which utilized data from a cohort of 2,760 women with IIH and 27,125 matched healthy controls from The Health Improvement Network (THIN) database. We employed innovative statistical models to adjust for the confounding effects of obesity, estimating the risk of ischemic stroke and cardiovascular disease attributable to IIH independent of obesity. Four distinct models were used to elucidate the complex interrelationships between IIH, obesity, and CVD risk.

**Results:** Our analysis revealed that IIH confers additional cardiovascular risk beyond that attributed to obesity alone. Risk ratios for various cardiovascular outcomes were consistently elevated across models comparing IIH patients to controls within the same obesity strata. A striking synergistic effect between IIH and obesity was observed, with the composite CVD risk reaching a risk ratio of 6.19 (95% CI: 4.58-8.36, p<0.001) in obese IIH patients compared to non-obese controls.

**Conclusions:** This study provides compelling evidence for a nuanced relationship between IIH, obesity, and cardiovascular risk. IIH appears to confer substantial cardiovascular risk independent of obesity, necessitating a paradigm shift in IIH management to encompass comprehensive cardiovascular risk mitigation. Further research is needed to elucidate the underlying mechanisms and develop targeted interventions for this unique patient population.

## 1. Introduction

Idiopathic intracranial hypertension (IIH) is a condition characterized by elevated intracranial pressure of unknown etiology, typically manifesting as papilledema with associated risks of visual loss and chronic disabling headache [1]. The incidence and economic burden of IIH are rising in parallel with global obesity trends [2]. While obesity is a well-established risk factor for IIH, with over 90% of patients being obese [3], the relationship between IIH and cardiovascular disease (CVD) risk remains poorly understood.

Adderley et al. conducted a retrospective case-control population-based matched controlled cohort study using 28 years of data from The Health Improvement Network (THIN) database in the United Kingdom, THIN database is a longitudinal primary care database containing anonymized electronic health records from over 17 million patients in the United Kingdom, provides researchers with comprehensive clinical data for epidemiological studies and healthcare research. [4]. Their study suggested that women with IIH have a two-fold increased risk of cardiovascular events compared to BMI-matched controls. However, the mechanisms underlying this elevated risk and the relative contributions of obesity versus IIH-specific factors remained unclear.

Building upon Adderley et al.’s [4] findings, our study aims to disentangle the effects of obesity and IIH on stroke risk specifically. Obesity is a known independent risk factor for stroke, with an average hazard ratio (HR) of 2.29 reported in large-scale evidence [5]. By adjusting for this obesity-related risk, we seek to isolate the potential contribution of IIH itself to stroke incidence.

Recent evidence suggests that IIH may be characterized by a unique profile of androgen excess affecting cerebrospinal fluid dynamics [6]. Androgen excess has been implicated as a key driver of increased cardiovascular risk in other conditions such as polycystic ovary syndrome [7, 8]. This raises the possibility that IIH may confer additional cardiometabolic risk beyond that attributable to obesity alone.

The prevalence and incidence of IIH have increased dramatically in recent years. Mollan et al. reported a tripling of IIH incidence in the UK between 2005 and 2017, from 2.3 to 7.8 per 100,000 person-years [2]. This rising disease burden underscores the urgent need to better understand and mitigate the long-term health risks associated with IIH.

Our study employs a novel methodological approach to simulate the predicted ischemic stroke and cardiovascular events in both IIH and control groups as if they had normal weight. By applying the established HR for obesity to adjust the observed events, we aim to estimate the risk of ischemic stroke and cardiovascular disease attributable to IIH independent of obesity. This approach assumes that the effect of obesity on stroke and cardiovascular disease risk remains constant over the study period and is independent of IIH status.

Understanding the relationship between IIH and their associated risks, independent of obesity, has important clinical implications. If IIH itself confers additional cardiovascular risk, it may warrant more aggressive management of modifiable risk factors and earlier implementation of preventive strategies in this patient population. Furthermore, elucidating the mechanisms underlying this potential association could reveal new therapeutic targets for reducing cardiovascular morbidity in IIH.

Our study aims to build upon the foundational work of Adderley et al. [4] to further investigate the complex interplay between IIH, obesity, and the associated risks. By employing innovative statistical methods to adjust for the confounding effects of obesity, we aim to provide crucial insights into the cardiovascular implications of IIH and inform evidence-based management strategies for this increasingly prevalent condition.

## 2. Methods

Building upon the foundational work of Adderley et al. [4], we conducted a retrospective analysis using data from their paper which was originally obtained through The Health Improvement Network (THIN), a large UK primary care database. Our study focused on women with IIH and matched controls, aiming to elucidate the independent effect of IIH on stroke and cardiovascular risks, distinct from the influence of obesity.

### 2.1. Study Population and Data Source

We utilized the cohort established by Adderley et al. [4], comprising 2,760 women with IIH and 27,125 matched controls. Participants were identified from THIN database records spanning January 1, 1990, to January 17, 2018. Controls were matched to IIH patients based on age, body mass index (BMI), and sex, with up to 10 controls per IIH case.

### 2.2. Outcome Measures

Our primary outcome of interest was the incidence of composite CVD, heart failure, ischemic heart disease (IHD), ischemic stroke, transient ischemic attack (TIA), hypertension, and type 2 diabetes mellitus. We extracted the relevant data from the corresponding paper, following the coding and identification methods described by Adderley et al [4].

### 2.3. Statistical Analysis

We extended the original analysis to estimate the independent effect of IIH on stroke and cardiovascular risks, accounting for the confounding effect of obesity. Our approach involved indirect standardization and adjustment with the application of a standardized morbidity ratio (SMR) approach [9-13], adapted to account for obesity as a confounding variable in relationship with IIH in women around the United Kingdom. To estimate the incidence of events in both the IIH and control cohorts under a hypothetical scenario of normal weight, we employed an adjustment method based on the average HR for obesity contributing to the event risk in women compared to healthy weight women in 13-year interval from the literature. This approach operates under the assumption that the HR remains constant throughout the 13-year study period and that the impact of obesity on the estimated events is independent of IIH status.

Initially, we calculated the observed HR for each event in the IIH group compared to the control group. Subsequently, we adjusted this observed HR by obesity HR to estimate the HR for IIH independent of obesity.

Based on the current evidence, the average estimated HR of obesity contributing to composite CVD is 2.89 [14-20]. For obesity, ischemic stroke, and TIA risk, it is estimated around HR= 1.72 [14, 17, 21-27]. For obesity and heart failure risk, it is estimated around HR= 2.61 [28-34]. For obesity and hypertension risk, it is estimated around HR= 2.09 [35-41]. For obesity and IHD risk, it is estimated around HR= 1.8 [14, 15, 17, 19, 21, 42, 43]. And for obesity and type 2 diabetes mellitus risk, it is estimated to be around HR= 4.0 [44-51].

We calculated the HR for each event in the IIH group compared to the control group through the following equation:

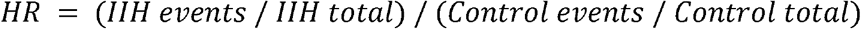

We then adjusted this observed HR by the established HRfor obesity in association with the potential risk to estimate the HR for IIH independent of obesity:

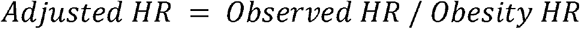

Using this adjusted HR, we predicted the number of events in both groups under normative weight conditions:

For the IIH group:

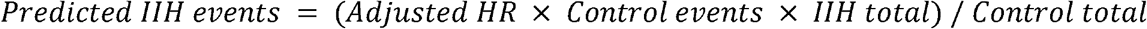

For the control group:

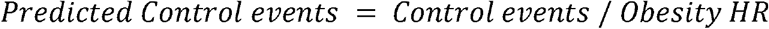

Using this adjusted HR, we then calculated the predicted number of events in both the IIH and control groups under the assumption of normal weight. This was accomplished by applying the adjusted HR to the control group event rate and scaling for the respective group sizes. For the control group, we divided the observed events by obesity HR to estimate events under normal weight conditions.

This method allows for a comparative analysis of events risk between IIH and control populations, while attempting to control the confounding effect of obesity. It provides insight into the potential independent risk associated with IIH and allows for estimation of event rates under hypothetical normal weight conditions.

### 2.4. Ethical Considerations

This study adhered to the ethical approval obtained by Adderley et al. [4] from the NHS South-East Multicenter Research Ethics Committee. We did not involve direct analysis of the dataset rather than building customized statistical modeling based on the provided data and metrics from Adderley et al. research paper [4].

## 3. Results

### 3.1. Baseline Characteristics

The original retrospective cohort study by Adderley et al. [4] encompassed 29,885 participants, stratified into 2,760 (9.2%) women with idiopathic intracranial hypertension (IIH) and 27,125 (90.8%) controls. The incident cohort comprised 48.2% and 46.7% of the IIH and control groups, respectively. Both cohorts were predominantly under 60 years of age (98.1% IIH, 95.2% control), with identical median ages of 32.1 years (IQR: 25.62-42.00 IIH, 25.71-42.06 control). Socioeconomic status, assessed via Townsend Deprivation Quintiles, showed a comparable distribution between groups, with a slight overrepresentation of controls in the least deprived quintiles. Smoking habits differed significantly: the IIH cohort exhibited higher rates of current smoking (30.8% vs 22.6%) and lower rates of non-smoking (46.5% vs 55.5%).

Anthropometric data revealed marginally higher median BMI in the IIH group (34.80, IQR: 29.30-40.30) compared to controls (34.30, IQR: 29.00-39.70). Notably, both groups demonstrated a high prevalence of obesity (BMI >30), affecting 62.6% and 60.9% of the IIH and control cohorts, respectively. Comorbidity profiles and pharmacological interventions showed distinct patterns. The IIH cohort demonstrated a higher prevalence of migraine (21.0% vs. 11.9%), hypertension (13.8% vs. 9.2%), and marginally increased rates of lipid-lowering medication use (6.5% vs. 5.8%). Furthermore, baseline cardiovascular morbidity was more pronounced in the IIH group, with elevated rates of ischemic heart disease (1.3% vs. 0.9%) and ischemic stroke/TIA (1.7% vs 0.7%). Interestingly, type 2 diabetes mellitus prevalence was slightly lower in the IIH cohort (4.7% vs. 5.2%), **Table 1**.

**Table 1.**
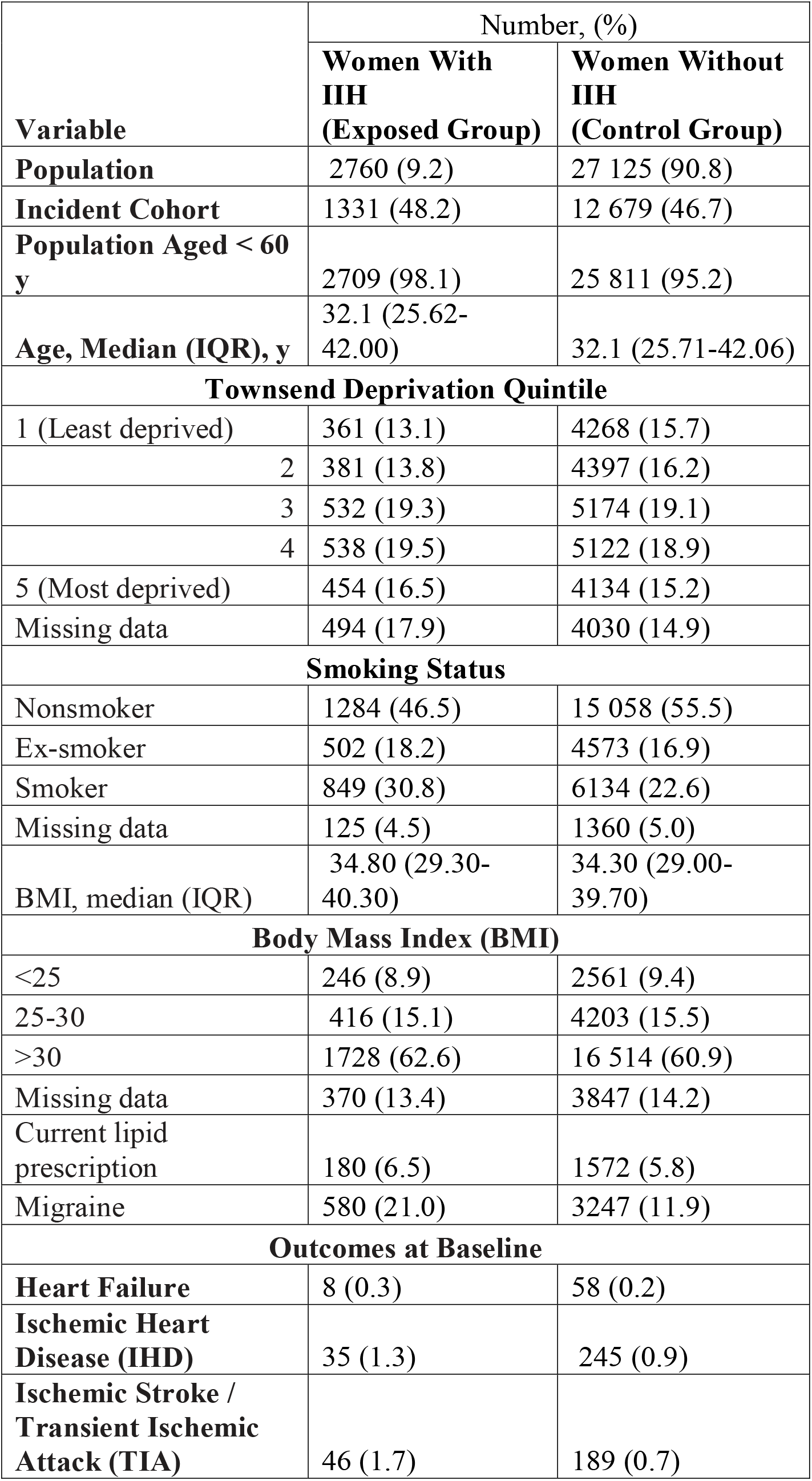
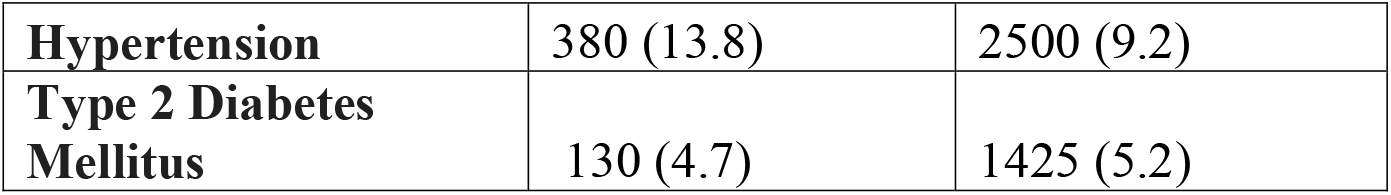
Baseline Characteristics of the Included Individuals in the Original Study.

| Variable | Number, (%) |  |
| --- | --- | --- |
|  | Women With IHH (Exposed Group) | Women Without IHH (Control Group) |
| Population | 2760 (9.2) | 27 125 (90.8) |
| Incident Cohort | 1331 (48.2) | 12 679 (46.7) |
| Population Aged < 60 y | 2709 (98.1) | 25 811 (95.2) |
| Age, Median (IQR), y | 32.1 (25.62-42.00) | 32.1 (25.71-42.06) |
| **Townsend Deprivation Quintile** |  |  |
| 1 (Least deprived) | 361 (13.1) | 4268 (15.7) |
| 2 | 381 (13.8) | 4397 (16.2) |
| 3 | 532 (19.3) | 5174 (19.1) |
| 4 | 538 (19.5) | 5122 (18.9) |
| 5 (Most deprived) | 454 (16.5) | 4134 (15.2) |
| Missing data | 494 (17.9) | 4030 (14.9) |
| **Smoking Status** |  |  |
| Nonsmoker | 1284 (46.5) | 15 058 (55.5) |
| Ex-smoker | 502 (18.2) | 4573 (16.9) |
| Smoker | 849 (30.8) | 6134 (22.6) |
| Missing data | 125 (4.5) | 1360 (5.0) |
| BMI, median (IQR) | 34.80 (29.30-40.30) | 34.30 (29.00-39.70) |
| **Body Mass Index (BMI)** |  |  |
| <25 | 246 (8.9) | 2561 (9.4) |
| 25-30 | 416 (15.1) | 4203 (15.5) |
| >30 | 1728 (62.6) | 16 514 (60.9) |
| Missing data | 370 (13.4) | 3847 (14.2) |
| Current lipid prescription | 180 (6.5) | 1572 (5.8) |
| Migraine | 580 (21.0) | 3247 (11.9) |
| **Outcomes at Baseline** |  |  |
| Heart Failure | 8 (0.3) | 58 (0.2) |
| Ischemic Heart Disease (IHD) | 35 (1.3) | 245 (0.9) |
| Ischemic Stroke / Transient Ischemic Attack (TIA) | 46 (1.7) | 189 (0.7) |
| **Hypertension** | 380 (13.8) | 2500 (9.2) |
| **Type 2 Diabetes Mellitus** | 130 (4.7) | 1425 (5.2) |

### 3.2. Statistical Analysis

In this analysis, we employed four distinct statistical models to elucidate the complex interrelationships between IIH, obesity, and cardiovascular disease (CVD) risk. These models were strategically designed to disentangle the individual and combined effects of IIH and obesity on cardiovascular outcomes.

Model 1 (Obese IIH vs Obese Control) was constructed to isolate the effect of IIH within an obese population, effectively controlling for the confounding factor of adiposity. Model 2 (Obese IIH vs Non-obese Control) provided a comprehensive view of the combined impact of IIH and obesity compared to individuals without either condition. Model 3 (Non-obese IIH vs Obese Control) offered a unique perspective, juxtaposing the cardiovascular risks associated with IIH in non-obese individuals against those attributed to obesity alone. Model 4 (Non-obese IIH vs. Non-obese Control) isolated the impact of IIH in a non-obese population, providing critical insights into the condition’s effects independent of obesity, **Table 2**.

**Table 2.**
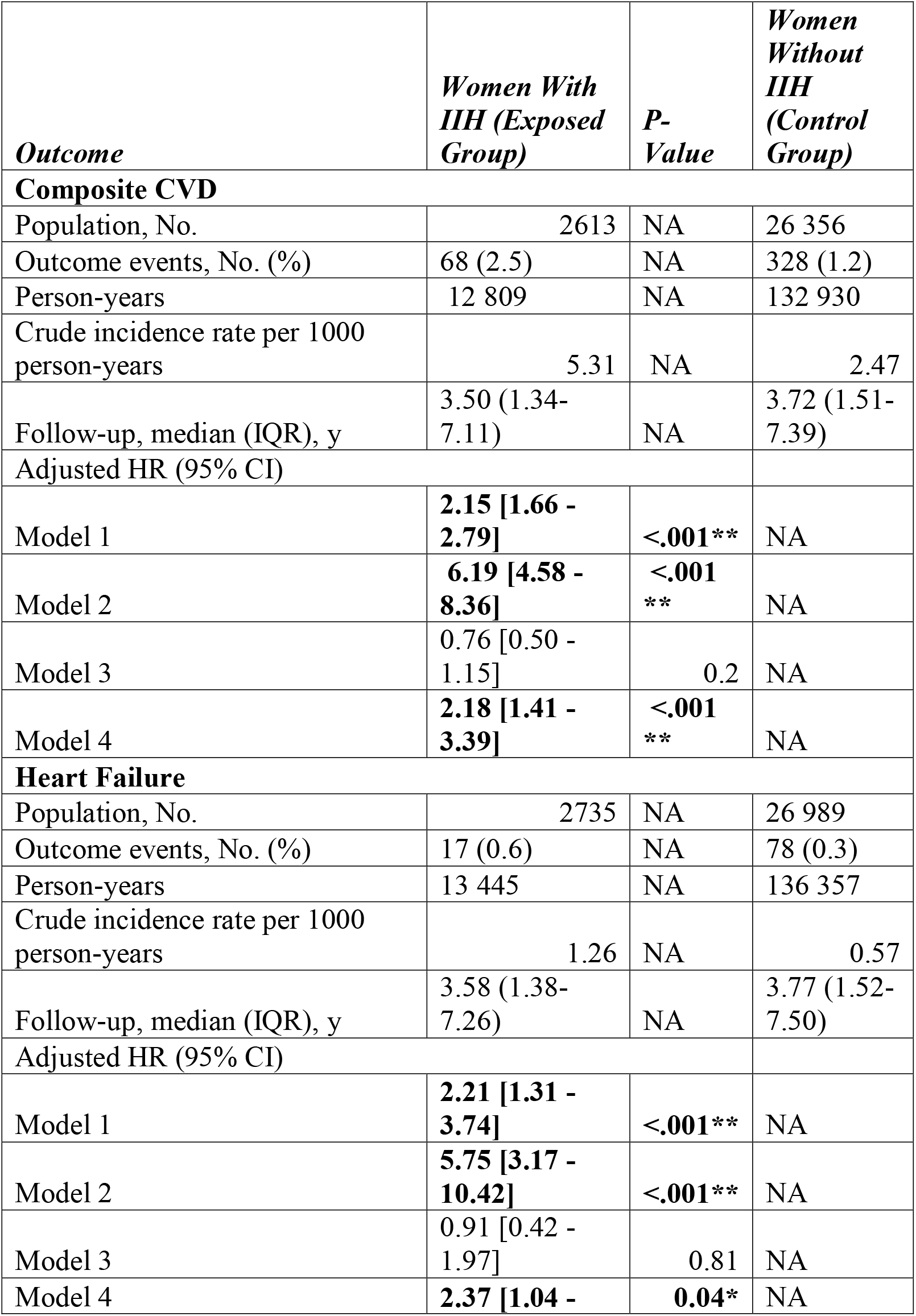
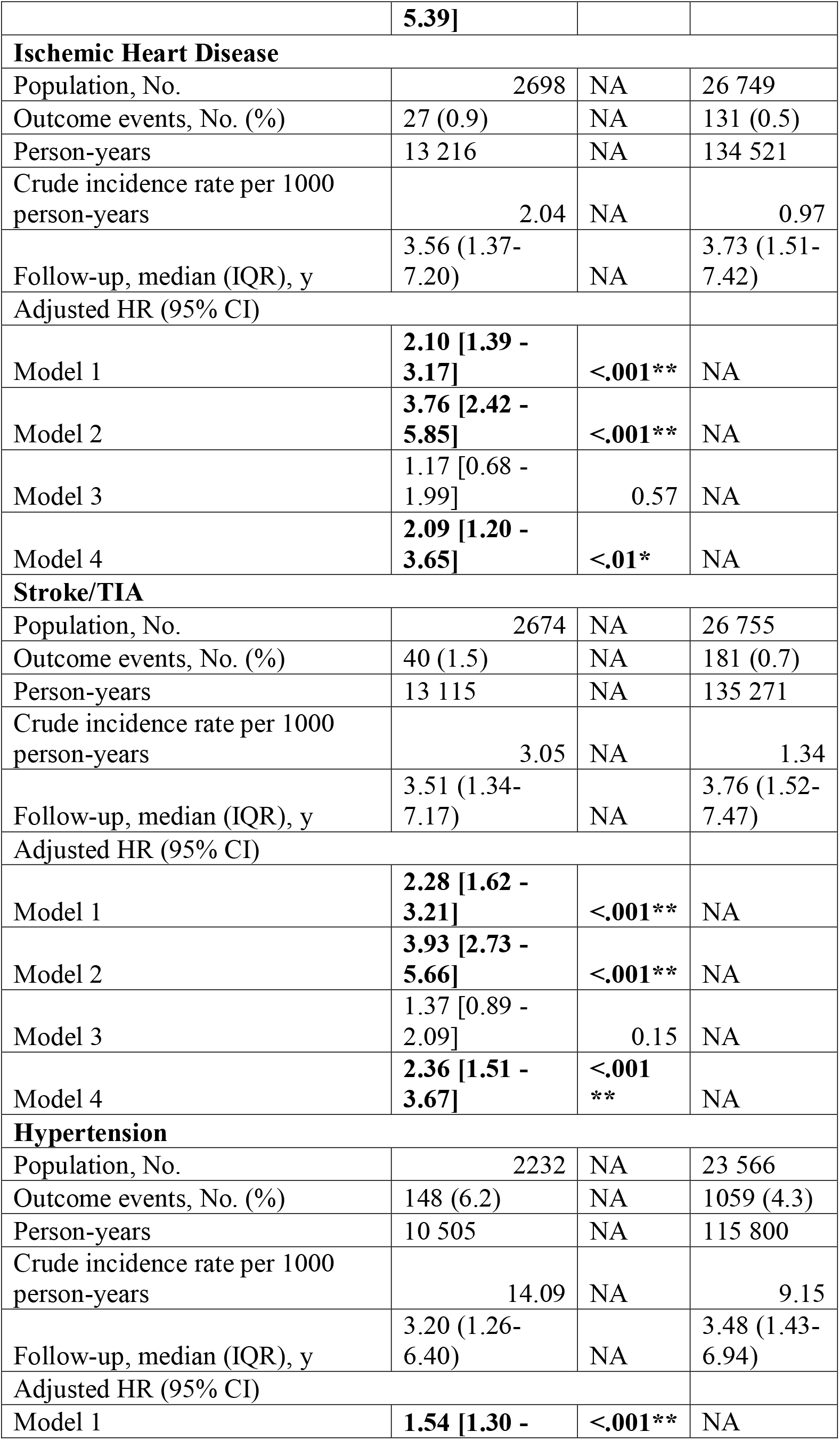
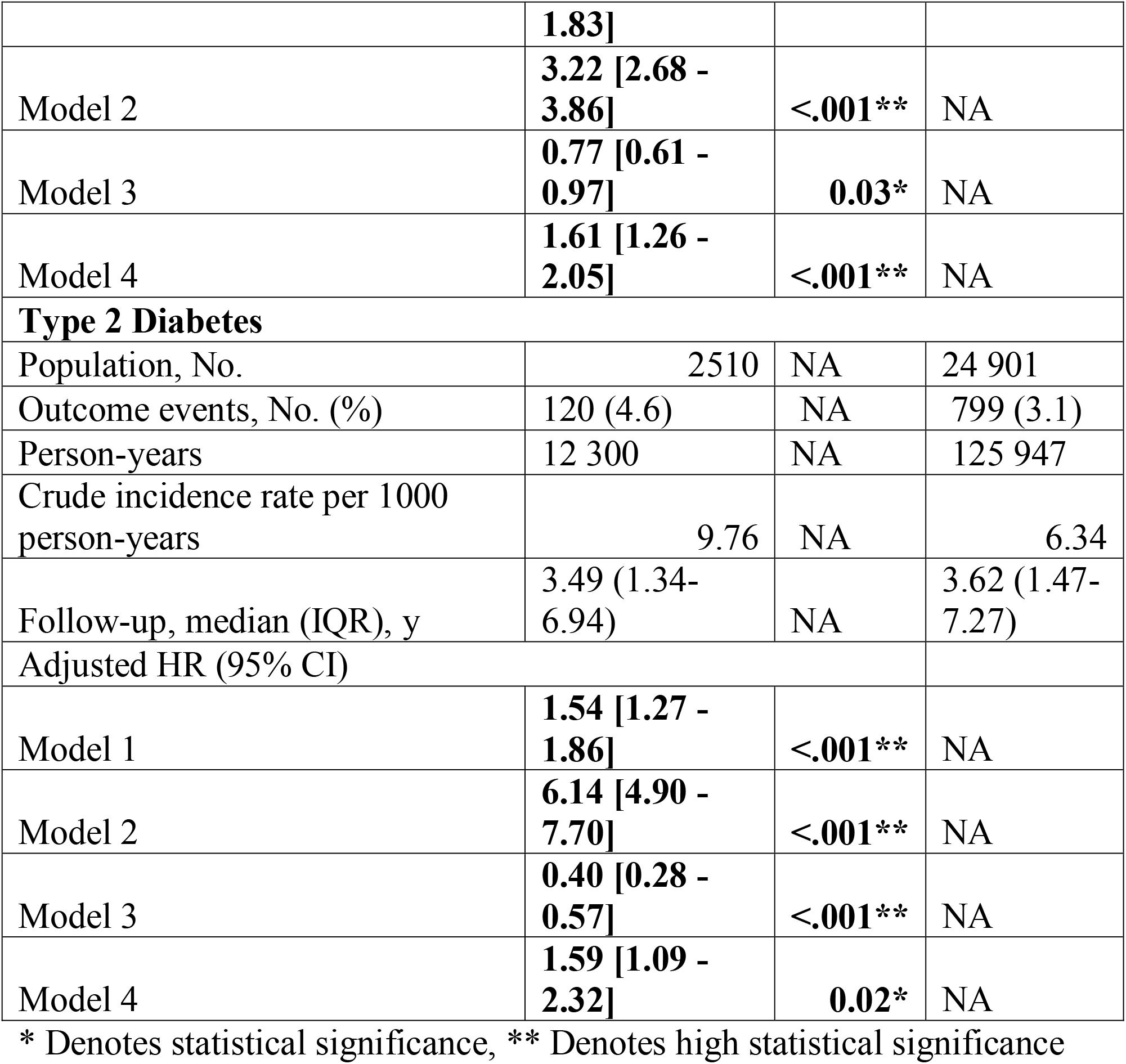
Risk Contribution Calculations According to Different Hazard Regression Models.

Our findings revealed a nuanced and clinically significant relationship between IIH, obesity, and cardiovascular risk. In Model 1 **Figure 1**, IIH was consistently associated with elevated risks across all measured outcomes. The risk ratios (RR) ranged from 1.54 (95% CI: 1.27-1.86, p<0.001) for type 2 diabetes mellitus to 2.28 (95% CI: 1.62-3.21, p<0.001) for stroke/TIA. This uniform pattern of risk elevation suggests that IIH confers additional cardiovascular risk beyond that attributed to obesity alone, a finding of relevance in clinical risk stratification.

**Figure 1.**
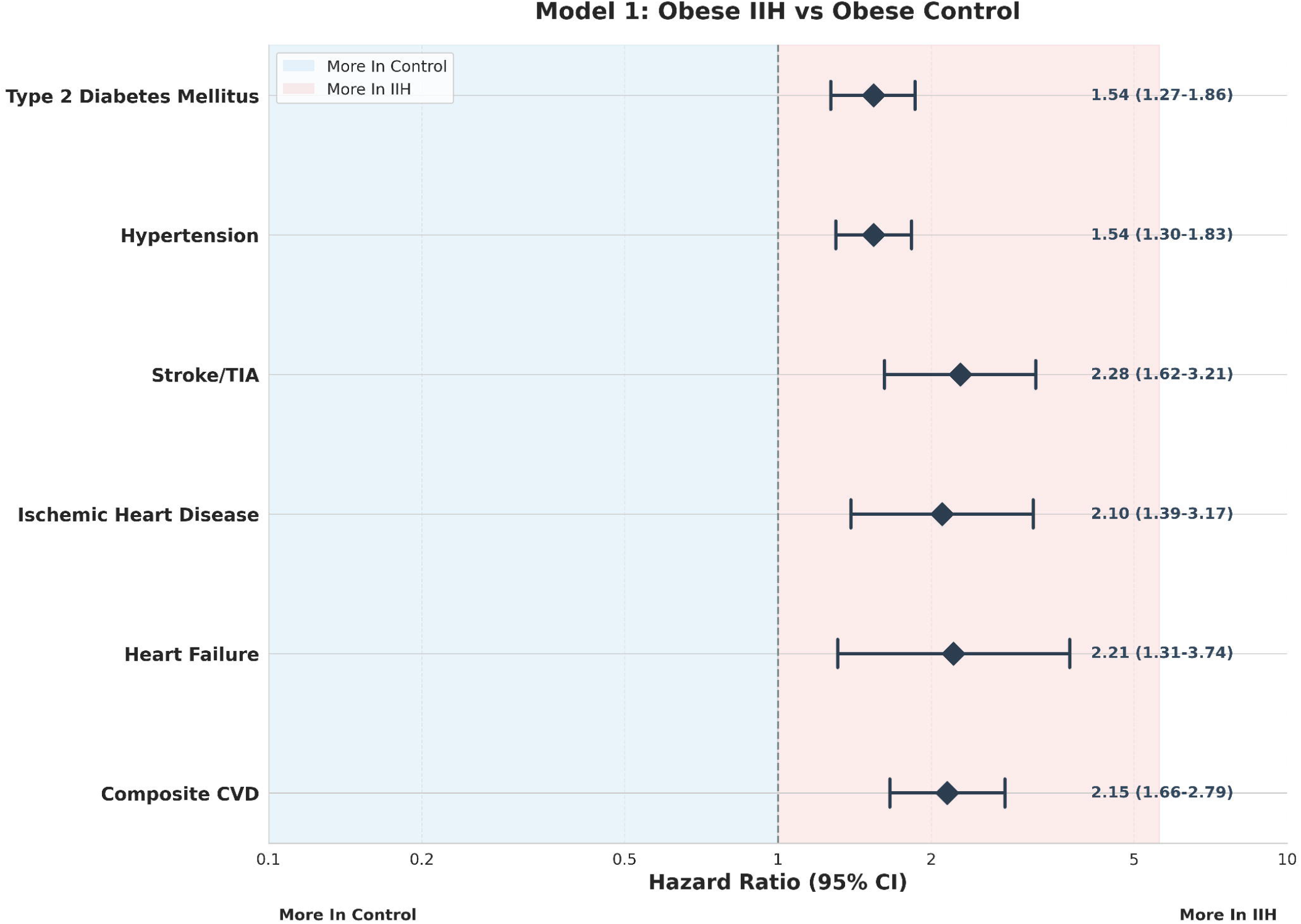
Model 1 – Obese IIH vs Obese Control Forest Plot.

Model 2, **Figure 2** demonstrated even more pronounced risk elevations, with the composite CVD risk reaching a striking RR of 6.19 (95% CI: 4.58-8.36, p<0.001). This marked increase suggests a potential synergistic effect between IIH and obesity on cardiovascular health, which may have significant implications for patient management and therapeutic interventions. Notably, the risk for heart failure in this model was particularly elevated (RR 5.75, 95% CI: 3.17-10.42, p<0.001), highlighting the need for vigilant cardiac monitoring in obese IIH patients.

**Figure 2.**
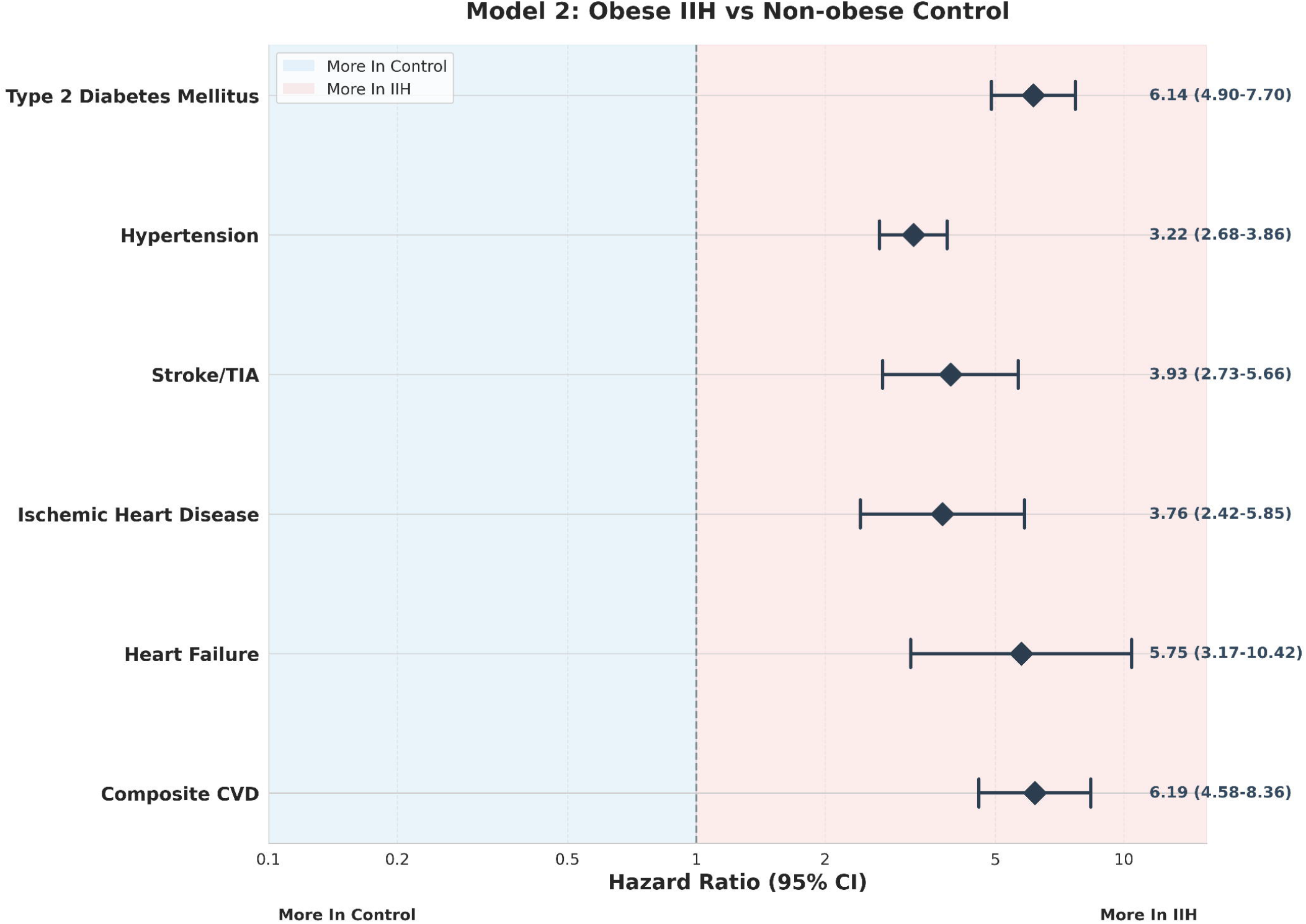
Model 2 – Obese IIH vs Non-Obese Control Forest Plot.

Interestingly, Model 3, **Figure 3**, presented a more complex picture. The non-significant risk ratios for most outcomes in this model suggest that non-obese individuals with IIH may not have significantly different cardiovascular risks compared to obese individuals without IIH. This finding underscores the profound impact of obesity on cardiovascular health, potentially rivaling or even overshadowing the effects of IIH in certain contexts. Of note in this model was the significantly reduced risk of type 2 diabetes mellitus in non-obese IIH patients compared to obese controls (RR 0.40, 95% CI: 0.28-0.57, p<0.001). This intriguing paradox may offer valuable insights into the underlying pathophysiology of both conditions and warrants further mechanistic investigation.

**Figure 3.**
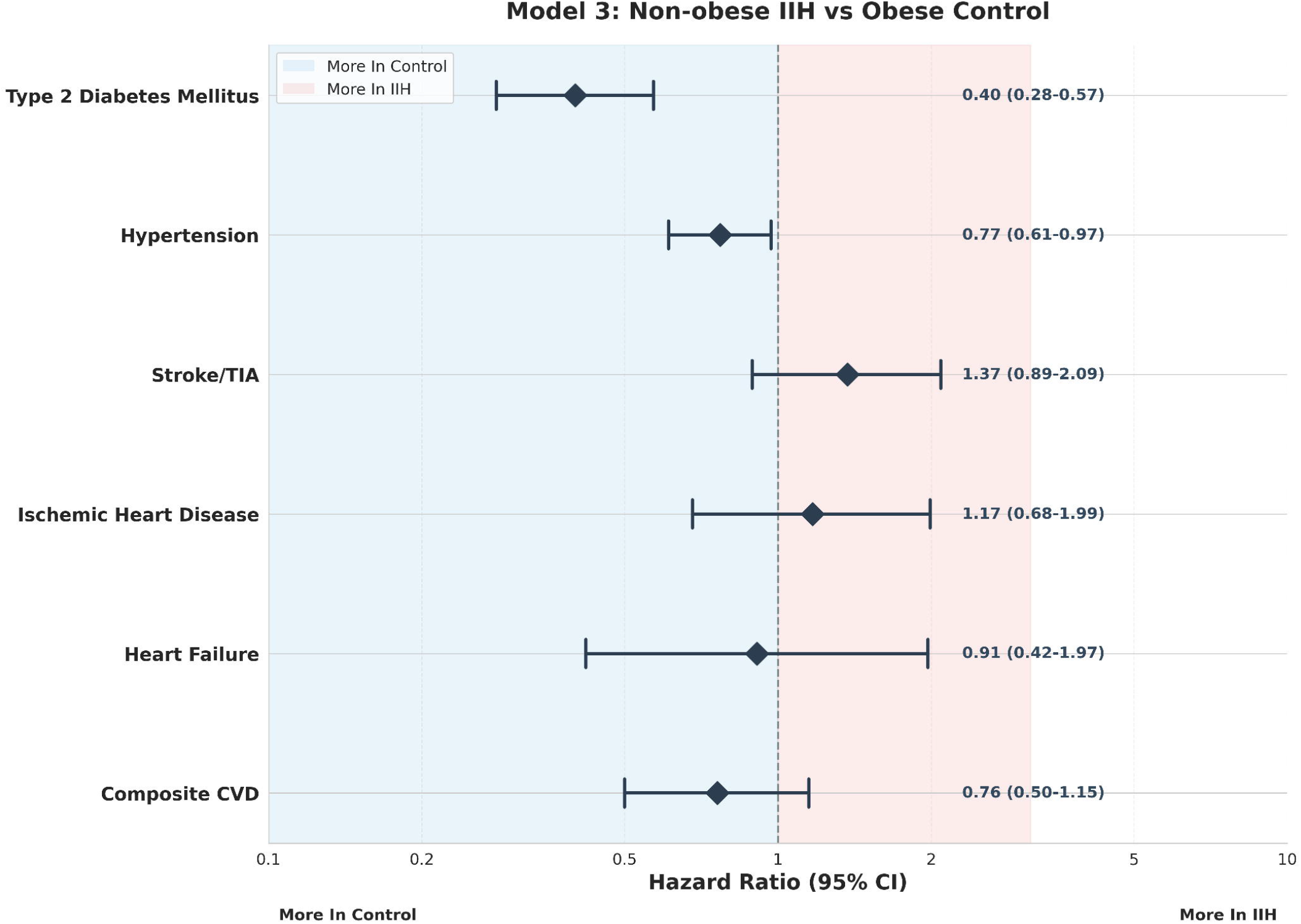
Model 3 – Non-Obese IIH vs Obese Control Forest Plot.

Model 4, **Figure 4** provided robust corroboration of IIH as an independent risk factor, with significant risk elevations observed across all outcomes in non-obese IIH patients compared to non-obese controls. The composite CVD risk in this model (RR 2.18, 95% CI: 1.41-3.39, p<0.001) closely mirrored that observed in Model 1, further supporting the notion that IIH confers cardiovascular risk independent of obesity status. This finding has important implications for the management of non-obese IIH patients, who may be at underappreciated cardiovascular risk.

**Figure 4.**
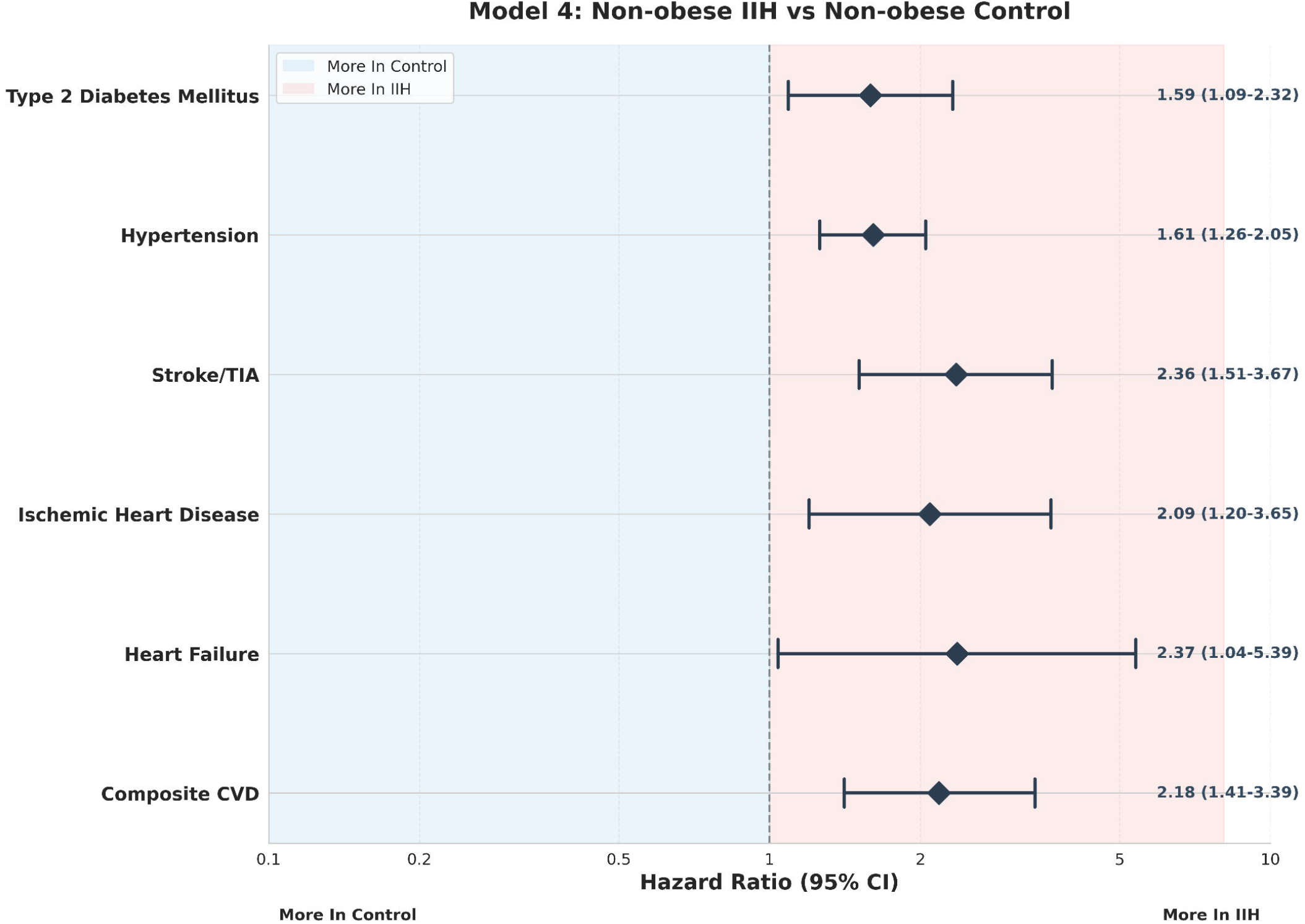
Model 4 – Non-Obese IIH vs Non-Obese Control Forest Plot.

Ranking the cardiovascular risks for IIH patients based on our data reveals the highest risk ratios in Model 2, with the following hierarchy: composite CVD (RR 6.19) > heart failure (RR 5.75) > stroke/TIA (RR 3.93) > ischemic heart disease (RR 3.76). This stratification underscores the critical importance of addressing both IIH and obesity in our highest-risk patients and may inform the development of targeted screening and intervention protocols. The data on type 2 diabetes mellitus warrant special consideration. The 6.14-fold increased risk (95% CI: 4.90-7.70, p<0.001) observed in obese IIH patients compared to non-obese controls (Model 2) is particularly striking. This marked elevation, coupled with the paradoxical risk reduction in non-obese IIH patients (Model 3), suggests a complex interplay between IIH, obesity, and metabolic dysfunction. These findings raise intriguing questions about potential shared pathophysiological mechanisms and may open new avenues for research into the neuroendocrine aspects of IIH. Hypertension, a known risk factor for both CVD and IIH progression, showed a consistent pattern of elevated risk across Models 1, 2, and 4. However, the reduced risk observed in Model 3 (RR 0.77, 95% CI: 0.61-0.97, p=0.03) adds another layer of complexity to our understanding of the relationship between IIH, obesity, and blood pressure regulation.

## 4. Discussion

Our obesity-adjusted risk analysis using indirect standardization analysis of the intricate relationship between IIH, obesity, and cardiovascular disease risk provides groundbreaking insights that not only corroborate but substantially extend the seminal work of Adderley et al [4]. By employing innovative statistical models to disentangle the complex interplay between IIH and obesity, we have unveiled a multifaceted paradigm of cardiovascular risk that challenges current understanding and opens new avenues for targeted interventions.

The consistent elevation of risk ratios across Models 1 and 4, which compare IIH patients to controls within the same obesity strata, strongly suggests a distinct pathophysiological process intrinsic to IIH that exacerbates cardiovascular vulnerability. This finding aligns with emerging research on the neuroendocrine and metabolic perturbations in IIH. Recent metabolomic profiling by O’Reilly MW et al [6]. revealed a unique signature of altered androgen metabolism in cerebrospinal fluid (CSF) of IIH patients, characterized by elevated levels of testosterone and androstenedione. This androgen excess may represent a crucial link between IIH and cardiovascular risk through multiple mechanisms, including vascular dysfunction, inflammatory modulation, and metabolic dysregulation. Duckles and Miller [52] demonstrated that testosterone could induce vasoconstriction through both genomic and non-genomic pathways, potentially contributing to hypertension and altered cerebrovascular autoregulation in IIH.

The chronic elevation of intracranial pressure (ICP) characteristic of IIH may have direct and indirect effects on cardiovascular function. Recent work by Wardlaw et al. [53] on the glymphatic system and intracranial fluid dynamics suggests that altered CSF flow and clearance in IIH may impair the removal of metabolic waste products from the brain. This accumulation of potentially toxic metabolites could exacerbate oxidative stress and vascular inflammation, contributing to the observed cardiovascular risk.

The striking risk elevations observed in Model 2 (Obese IIH vs Non-obese Control) reveal a synergistic interaction between IIH and obesity that amplifies cardiovascular risk beyond the sum of their individual effects. This synergy likely arises from the convergence of multiple pathophysiological processes, including adipokine dysregulation, neuroendocrine activation, and hemodynamic alterations. Recent work by Hornby et al. [54] demonstrates that IIH patients exhibit a distinct adipokine signature, with particularly elevated CSF leptin levels. The combination of systemic and central adipokine dysregulation may create a uniquely pro-inflammatory and pro-thrombotic state. Moreover, the evidence by Markey K et al. [55] suggests that IIH patients may have altered cortisol metabolism, potentially exacerbating the metabolic and cardiovascular consequences of obesity related Hypothalamic-Pituitary-Adrenal axis dysfunction.

The paradoxical findings regarding type 2 diabetes risk in our study—elevated in obese IIH patients but reduced in non-obese IIH patients compared to obese controls—challenge our current understanding of metabolic risk in IIH. This observation may be explained by the concept of “metabolic flexibility” proposed by Goodpaster and Sparks [56]. In non-obese IIH patients, the altered androgen metabolism and potential changes in adipose tissue function may confer a degree of metabolic protection. The evidence by Mariniello et al. [57] on androgen effects on adipose tissue suggests that certain androgen profiles can enhance insulin sensitivity and improve glucose uptake in adipocytes. The specific androgen milieu in IIH may thus have differential effects depending on the overall metabolic context. Conversely, in obese IIH patients, this potential metabolic benefit may be overwhelmed by the profound insulin resistance and chronic inflammation associated with obesity. The interaction between obesity-related metabolic dysfunction and IIH-specific neuroendocrine perturbations may create a “perfect storm” for accelerated progression to type 2 diabetes [57].

Our findings necessitate a paradigm shift in the approach to cardiovascular risk management in IIH patients. We propose a multi-tiered strategy that includes enhanced risk stratification, targeted interventions, personalized metabolic management, and neuroendocrine modulation. The development of IIH-specific cardiovascular risk calculators that incorporate novel biomarkers such as CSF androgen levels, adipokine profiles, and measures of intracranial pressure dynamics could significantly improve risk assessment in this population. Exploration of IIH-specific pharmacological interventions that address the unique pathophysiology of cardiovascular risk in this population is warranted. For example, the potential use of selective androgen receptor modulators (SARMs) to mitigate the adverse cardiovascular effects of androgen excess while preserving potential metabolic benefits merits investigation.

Future research directions should include longitudinal studies employing advanced imaging techniques to elucidate the temporal relationship between IIH onset, progression, and cardiovascular remodeling. Multi-omics approaches integrating genomics, transcriptomics, and metabolomics could unravel the molecular mechanisms underlying the observed synergy between IIH and obesity in cardiovascular risk. Interventional trials exploring the cardiovascular impact of IIH-specific treatments, including the potential cardioprotective effects of CSF diversion procedures or novel pharmacological agents targeting ICP regulation, are crucial. Additionally, investigation of sex-specific aspects of cardiovascular risk in IIH is essential, given the strong female predominance of the condition and the potential interaction with sex hormones.

The findings from our study reveal a complex, multifaceted relationship between IIH, obesity, and cardiovascular risk that challenges existing paradigms and opens new frontiers in personalized medicine. The independent risk conferred by IIH, the synergistic effects with obesity, and the paradoxical metabolic findings underscore the need for a nuanced, mechanism-based approach to cardiovascular risk management in this unique patient population. As we continue to unravel the intricate pathophysiology of IIH, we move closer to developing targeted interventions that may not only alleviate the neurological symptoms of the condition but also mitigate its long-term cardiovascular consequences. The implications of our findings extend beyond IIH, offering potential insights into the broader interplay between neuroendocrine function, metabolic regulation, and cardiovascular health. The methodology of our paper has several limitations, at first the approach assumes that the HR and the values provided from the original data and the HR for obesity remains constant over the 13-year period and its applicable to both the IIH group and control group. Secondly, it assumes that the effect of obesity on the events is independent of IIH status in each patient. Thirdly, the predicted events are based on the average HR for obesity from the current literature, which may not be fully representative of the study population in larger populations or another cohort. Also, the adjusted for IIH independent from obesity should be interpreted with caution, as it is an estimation based on the available data and assumptions. To further validate the findings, it would be better to perform tailored individual-level data analysis based on BMI subgroup analysis and sensitivity tests for IIH patients and counting for other potential cofounding variables in the cohort. Additionally, conducting a prospective study that directly compares IIH patients with normal weight controls would provide more comprehensive evidence for the independent effect of IIH on the proposed events.

## 5. Conclusions

Our study provides compelling evidence for a nuanced relationship between idiopathic intracranial hypertension (IIH), obesity, and cardiovascular risk. Through innovative statistical modeling, we have disentangled the independent effects of IIH from those of obesity, revealing a complex interplay with significant clinical implications. Our findings demonstrate that IIH confers substantial cardiovascular risk beyond that attributable to obesity alone. The consistent elevation of risk ratios across obesity-stratified models suggests an intrinsic pathophysiological process in IIH that exacerbates cardiovascular vulnerability. This aligns with emerging research on neuroendocrine perturbations in IIH, particularly the unique androgen metabolic signature recently identified in cerebrospinal fluid. The striking synergy between IIH and obesity in amplifying cardiovascular risk, as evidenced by our Model 2 results, underscores the critical importance of addressing both conditions concurrently. This synergistic effect likely arises from the convergence of multiple pathways, including adipokine dysregulation, altered intracranial fluid dynamics, and systemic metabolic dysfunction. Our paradoxical findings regarding type 2 diabetes risk—elevated in obese IIH patients but reduced in non-obese IIH patients compared to obese controls—challenge current understanding and suggest a complex metabolic phenotype in IIH. This observation may reflect the concept of “metabolic flexibility” and warrants further investigation into the differential effects of IIH-specific neuroendocrine changes in varying metabolic contexts. These results necessitate a paradigm shift in IIH management, calling for a multi-faceted approach that extends beyond intracranial pressure control to encompass comprehensive cardiovascular risk mitigation. We propose the development of IIH-specific cardiovascular risk calculators incorporating novel biomarkers and the exploration of targeted interventions addressing the unique pathophysiology of cardiovascular risk in this population.

Our study’s limitations, including assumptions about constant hazard ratios and obesity effects, underscore the need for further research. Prospective studies directly comparing IIH patients with normal-weight controls and employing advanced multi-omics approaches will be crucial to validating and extending our findings.

## IRB Approval

N/A

## Ethical Approvals

N/A

## Consent for Participation

N/A

## Consent to Publish

N/A

## Data Availability Statement

N/A

